# EVALUATON OF INFLAMMATION WITH CRP, IL-1, PRO-BNP, LEPTIN AND PETRAXIN -3 LEVELS IN OBESITY-RELATED HYPERTENSION

**DOI:** 10.1101/2024.08.16.24312093

**Authors:** Gürkan Karakuş, Cemşit Karakurt, Özlem Elkıran, Mehmet Öncül, Serdar Akın Maraş, Yılmaz Tabel, Emine Çamtosun, Çağatay Taşkapan, Harika Gözde Gözükara Bağ

**Affiliations:** İnönü University Faculty of Medicine, Department of Pediatrics, Malatya, Türkiye; İnönü University Faculty of Medicine, Department of Pediatric Cardiology, Malatya, Türkiye; İnönü University Faculty of Medicine, Department of Pediatric Nephrology, Malatya, Türkiye; İnönü University Faculty of Medicine, Department of Pediatric Endocrinology and Metabolism, Malatya, Türkiye; İnönü University Faculty of Medicine, Department of Biochemistry, Malatya, Türkiye; İnönü University Faculty of Medicine, Department of Bisostatistics, Malatya, Türkiye

**Keywords:** Obesity, hypertension, inflammation, ambulatory blood pressure monitoring, echocardiography

## Abstract

**Purpose:** The aim of study is to determine the relationship between the inflammation and the severity of hypertension with the inflammation biomarkers; CRP, IL-1, proBNP, leptin, PTX-3 levels in children with obesity-related hypertension.

**Material and Methods:** In our study, thirty patients (20 girls, 10 boys) who were admitted to İnönü University Turgut Özal Tıp Merkezi Department of Pediatrics between 2021-2022; between the ages of 2-17 and who were followed up in Pediatric Endocrinology, Pediatric Nephrology and Pediatric Cardiology clinics for obesity-related hypertension and 30 healthy children (22 girls, 8 boys) in the same age group were included. Clinical and demographic characteristics of all children; Age, gender, anthropometric measurements, blood pressure, 24-hour ambulatory blood pressure measurements echocardiographic findings were recorded. Left ventricular systolic and diastolic diameters, volumes and wall thickness measured by echocardiography and proportioned to body surface area. After the echocardiographic evaluation, a total of 10 cc blood was drawn from the individuals and patients in the control group and IL-1, CRP, Leptin, Pro-BNP, PTX-3 levels were measured. Statistical analysis was performed using SPSS (Statistical Program in Social Sciences) 22.0 software program

**Results:** Systolic blood pressure, diastolic blood pressure, height, weight, body mass index were statistically significantly higher in the patient group. On echocardiographic evaluation, interventricular diastolic diameter (IVSd), left ventricular diastolic diameter (LVIDd), left ventricular posterior wall diastolic (LVPWd), interventricular systolic diameter (IVSs), left ventricular systolic diameter (LVIDs) and left ventricular diastolic diameter (LVIDd), left ventricular posterior wall systolic diameter (LVPWs) were statistically significantly higher in the patient group. LVs mass values, systolic blood pressure, diastolic blood pressure, height, weight, body mass index, IVSd, LVIDd, LVPWd, IVSs, LVIDs, LVPWs, LVs massvalues were statistically significantly higher in the patient group. In our study, when the inflammation variables in the patient and control groups were compared, leptin and CRP levels were found significantly higher in the obesity group (p:0,041). We found that there was a moderate positive correlation between leptin and systolic blood pressure in healthy children, moderately strong correlations between Pro-BNP and systolic blood pressure in the negative corelation, and a weak positive correlation between leptin and diastolic blood pressure in obesity group. When the correlations between inflammation parameters and blood pressure findings were examined in our study, positive and moderate (p:0,019 r:0,425) correlation was found between leptin and systolic blood pressure in healthy children, while negative moderate (p:0,001 r:-0,566) correlation between Pro-BNP and systolic blood pressure and positive weak correlation between leptin and diastolic blood pressure (p:0,036 r:0,384) were found in obese and hypertensive children. Leptin, systolic blood pressure, diastolic blood pressure, height, weight and body mass index, IVSd, LVIDd, LVPWd, IVSs, LVIDs, LVPWs and LVs mass values were found to be statistically significantly high in obese patients who were required anti-hypertensive treatment.

**Conclusion:** The importance of inflammatory markers in the early diagnosis of obesity-related hypertension may guide the development of new therapeutic strategies to treat target organ damage by reducing the morbidity and mortality that can be caused by hypertension. . We think that the high levels of inlammation markers in obese patients may be a guide in the severity of hypertension, in the prevention of organ damage secondary to hypertension, in reversing inflammation and hypertension with more stringent lifestyle changes, diet and exercise.

However, prospective studies which compare pre- and post-treatment inflammation in children are needed to evaluate the effects of inflammation and its effects on prognosis.

## INTRODUCTION

Obesity describes the increase in body fat over the normal size of the body. The most commonly used method in obesity screening is body mass index (BMI) calculation (1). In children and youngs up to 20 years of age, obesity is defined as a body mass index (BMI) of ≥ 95. age and gender specific percentile. Being overweight is defined as BMI between ≥ 85. and <95. percentile. In children, severe obesity is defined as 120% of BMI ≥ 95. percentile (corresponding to 99. percentile BMI) or BMI ≥ 35 kg/m^2^ (corresponding to the cut-off point for Class II obesity in adults). After the age of 20, adult limit values for overweight and obesity apply (BMI between 25 and 30 kg/m^2^ and BMI ≥30 kg/m^2^, respectively).

In studies conducted on adults, when compared with adults with normal BMI, the risk of hypertension was found to increase up to 4.8 times with high BMI rate in adults with Class III obesity (BMI≥40 kg/m^2^) (2). Mainly two basic hormonal mechanisms are emphasized in obesity-related hypertension. The first mechanism is hyperinsulinemia, occurs as a result of peripheral insulin resistance; the second mechanism is hyperleptinemia, occurs due to an increase in fat tissue mass (3). Obesity is a common vascular and systemic inflammatory condition. Obesity induced proinflammatory cytokines and oxidative stress increase peripheral vascular resistance by causing vascular endothelial dysfunction and disruption of local vasodilator responses (4). Insulin resistance, low adiponectin level, increase in plasma leptin level, increased plasma glucose and free fat acids are important indicators of inflammation (5). Since changes in circulating cytokine levels and acute phase reactants are defined in obese individuals, it is defined as a condition with chronic low-grade systemic inflammation (6) (7). In a large number of studies conducted so far, obese children have been compared with healthy children in terms of left ventricular measurements and left ventricular mass was found to increase significantly in obese children when compared with healthy children (8) (9). The aim of this study is to determine the relationship between the inflammation and the severity of hypertension with the inflammation biomarkers; CRP, IL-1, proBNP, leptin, PTX-3 levels in children with obesity-related hypertension.

## MATERIAL and METHOD

This cross sectional prospective study was conducted with children between the ages of 2 and 17 who were followed for obesity-related hypertension at İnönü University Turgut Özal Tıp Merkezi Department of Child Health and Diseases Pediatric Endocrinology, Pediatric Nephrology and Pediatric Cardiology Clinics between 2021 and 2022. Our study was conducted with a total of 60 pediatric patients, 30 in the patient group and 30 in the control group. Our exclusion criterias were presence of major cardiac problem in the echocardiographic evaluation, being younger than 2 years of age and older than 17 years of age, having systemic diseases other than obesity and hypertension. Before study ethical permission was obtained from İnönü University Faculty of Medicine Health Sciences Clinical Research Ethics Committee and consent was obtained the individuals/parents who participated in the study.

After the participants rest at least 5 minutes in the clinic, their blood pressure was measured manually three times from the right arm with a sphygmomanometer by expert medical personel in sitting position.Measurement was taken at least 3 minutes between intervals and mean of the 3 measurements was taken. The arm was taken to keep at heart level during measurements. When necessary, the patients were evaluated with ambulatory blood pressure measurement (ABPM).

2D and color Doppler echocardiographic evaluation was performed by the same pediatric cardiologist according the recommendations of “*American Society of Echocardiography*”, by using with Vivid Pro–7 GE (GE Healthcare, Florida, USA). After structural heart diseases were excluded with 2D and color Doppler echocardiographic evaluation, left ventricular diameter, wall thickness and functions were evaluated by measuring with M mode in the parasternal long axis. Left ventricular measurements were made according to the recommendations of American Society of Echocardiography as interventricular septum diastolic thickness (IVSd), interventricular septum systolic thickness (IVSs), left ventricular diastolic diameter (LVIDd), left ventricular systolic diameter (LVIDs), left ventricular posterior wall diastolic thickness (LVPWd), left ventricular posterior wall systolic thickness (LVPWs), left ventricular diastolic mass (LVd mass), left ventricular systolic mass (LVs mass), left ventricular diastolic mass thickness (LVd mass-ASE), left ventricular systolic mass thickness (LVs mass-ASE). Left ventricular diameter, volume parameters and wall thickness parameters measured by echocardiography were proportioned to body surface area (m^2^).

After echocardiographic evaluation, a total of 10 cc blood was taken from the individuals in the control group and the patients into 2 biochemistry tubes (to measure IL-1, CRP, Leptin, Pro-BNP, PTX-3 levels) and the serums separated by centrifugation were stored at −80 °C.

SPSS 22 program was used in data analysis. Kolmogorov Smirnov test was used for normality distribution test. Parametric tests were used in the analysis of normally distributed data, while non-parametric tests were used in the analysis of non-normally distributed data. Mann Whitney U test, student t test, Spearman and Pearson correlation analysis were used in data analysis. p<0.05 value was considered as statistically significant in all tests.

Anthropometric measurements, blood pressure measurements and echocardiography measurements of the patients were also recorded. Inflammation parameters and echocardiography results were compared between the two groups.

## RESULTS

This cross sectional prospective study was conducted with children between the ages of 2 and 17 who were followed for obesity-related hypertension at İnönü University Turgut Özal Medical Center Department of Child Health and Diseases Pediatric Endocrinology, Pediatric Nephrology and Pediatric Cardiology Clinics between 2021 and 2022.

Mean age of the 30 obese and hypertensive patients included in the study was found as 13.86±3.38 years, while the mean age of the 30 individuals in the control group was found as 12.40±2.47 years. Demographic characteristics and anthropometric measurements of the patient and the control group are shown in Table 1.

**Table 1.** Demographic characteristics and anthropometric measurements of the patient and the control group.

|  | GROUP |  |  |
| --- | --- | --- | --- |
|  | Patient n:30 | Healthy n:30 | p |
| **Age (years)** | 13.86±3.38 | 12.40±2.47 | 0.016 |
| **Gender** |  |  |  |
| **Male** | 10(33.3) | 8(26.7) | 0.778 |
| **Female** | 20(66.7) | 22(73.3) |  |
| **Height (cm)** | 160,90±14,53 | 155,64±12,76 | 0.042 |
| **Weight (kg)** | 83,38±25,39 | 48,47±13,03 | <0.001 |
| **Body Mass Index (kg/m<sup>2</sup>)** | 31,24±6,05 | 19,61±2,85 | <0.001 |
| **Body Mass Index (percentile)** | 98,89±1,10 | 42,81±28,38 | <0.001 |
| **Systolic Blood Pressure (mmHg)** | 141,73±12,15 | 110,43±4,50 | <0.001 |
| **Diastolic Blood Pressure (mmHg)** | 81,50±11,95 | 62,93±4,87 | <0.001 |

On the echocardiographic evualation, IVSd: intraventricular semptum diastolic diameter, IVSs: intraventricular semptum systolic diameter, LVIDd: left ventricular diastolic diameter, LVIDs: left ventricular systolic diameter, LVPWd: left ventricular free wall diastolic diameter, LVPWs: left ventricular free wall diastolic diameter were found to be statistically higher in the patient group (Table 2).

**Table 2.** Comparison of echocardiography findings of the children in the patient and control group.

| Variable | GROUP |  |  |  |  |
| --- | --- | --- | --- | --- | --- |
|  | Patient n:30 |  | Control n:30 |  | p |
|  | X±S.S. | Median | X±S.S. | Median |  |
| IV Sd | 0,60±0,13 | 0,56 | 0,49±0,12 | 0,47 | **0,004** |
| LV IDd | 2,98±0,42 | 2,89 | 2,35±0,37 | 2,35 | **<0,001** |
| LV PWd | 0,56±0,14 | 0,53 | 0,48±0,11 | 0,49 | **0,046** |
| IV Ss | 0,76±0,14 | 0,74 | 0,64±0,11 | 0,62 | **<0,001** |
| LV IDs | 1,83±0,27 | 1,80 | 1,35±0,28 | 1,32 | **<0,001** |
| LV PWs | 0,92±0,25 | 0,85 | 0,75±0,14 | 0,74 | **0,002** |
| LVd mass | 86,74±33,32 | 79,41 | 80,12±16,31 | 78,42 | 0,871 |
| LVs mass | 66,23±22,86 | 60,86 | 59,84±15,68 | 60,17 | 0,712 |
| LVd mass AS | 75,96±26,39 | 69,33 | 72,26±12,92 | 71,31 | 0,965 |
| LVs mass AS | 61,23±13,98 | 57,34 | 57,74±17,83 | 53,37 | 0,169 |

When the inflammation parameters were compared, CRP and leptin levels were found to be statistically significant in the patient group. No statistically significant difference was found between groups in terms of Pentraxin-3, IL-1 and Pro-BNP values (Table 3).

**Table 3.**
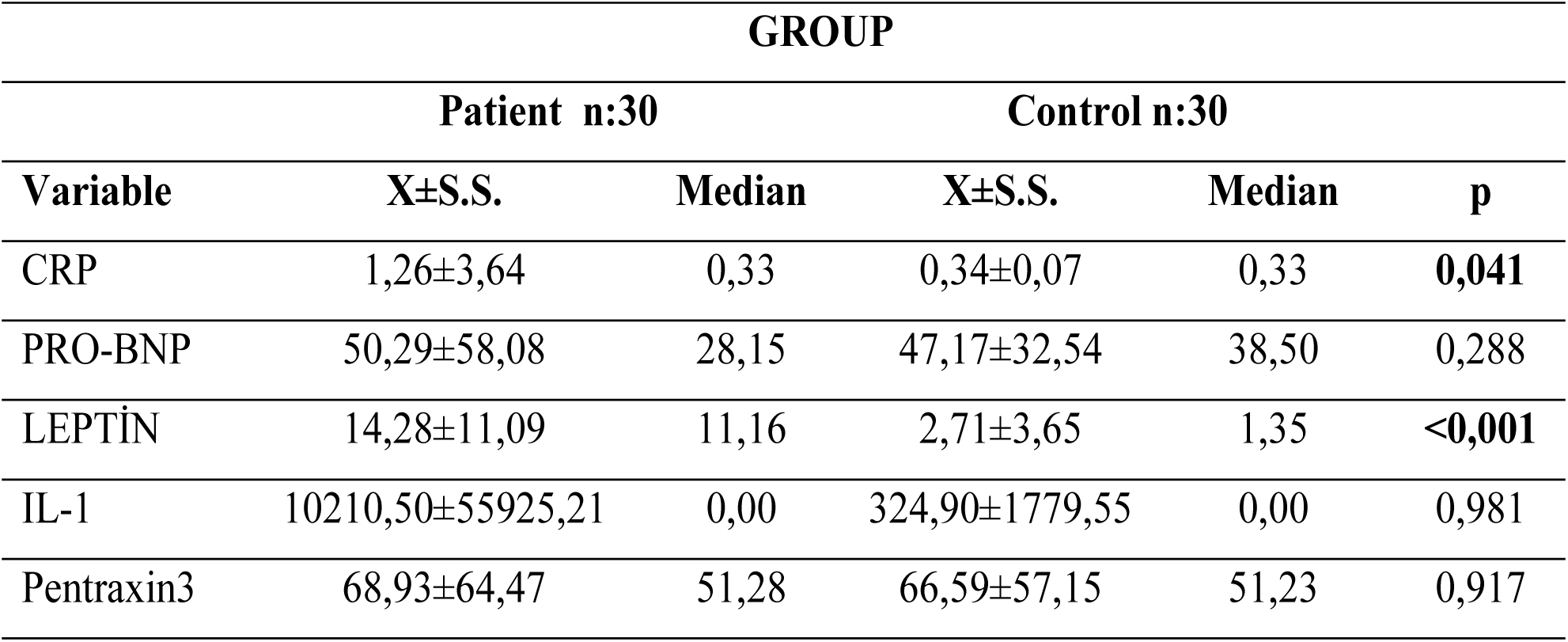
Comparison of inflammation parameters in terms of groups.

| **GROUP** |  |  |  |  |  |
| --- | --- | --- | --- | --- | --- |
| **Patient n:30** |  |  | **Control n:30** |  |  |
| **Variable** | **X±S.S.** | **Median** | **X±S.S.** | **Median** | **p** |
| CRP | 1,26±3,64 | 0,33 | 0,34±0,07 | 0,33 | **0,041** |
| PRO-BNP | 50,29±58,08 | 28,15 | 47,17±32,54 | 38,50 | 0,288 |
| LEPTİN | 14,28±11,09 | 11,16 | 2,71±3,65 | 1,35 | **<0,001** |
| IL-1 | 10210,50±55925,21 | 0,00 | 324,90±1779,55 | 0,00 | 0,981 |
| Pentraxin3 | 68,93±64,47 | 51,28 | 66,59±57,15 | 51,23 | 0,917 |

When the correlations between Pentraxin-3 and CRP and echocardiography results were examined in the patient and control group, no significant correlation was found (table 4). Statisticaly correlation was found between Pro-BNP and echocardiography results in the patient and control group. Negative and moderate correlation was found between **LVd mass:** left ventricular diastolic mass and **LVs mass:** left ventricular systolic mass and Pro-BNP (table 4) (p:0,02 r:-0,392 and p:0,006 r:-0,492)

**Table 4.** Correlation between inflammation parameters and left ventricular diameter, wall thicknesses and mass measurements.

|  |  | PATIENT |  |  |  | CONTROL |  |  |  |
| --- | --- | --- | --- | --- | --- | --- | --- | --- | --- |
|  |  | CRP | ProBNP | LEPTIN | PTX-3 | CRP | ProBNP | LEPTIN | PTX-3 |
| IV Sd | R | -0,326 | 0,023 | -0,014 | 0,085 | 0,054 | 0,087 | 0,018 | -0,133 |
|  | P | 0,079 | 0,903 | 0,942 | 0,654 | 0,778 | 0,649 | 0,923 | 0,483 |
| LV IDd | R | -0,136 | -0,290 | -0,441 | -0,180 | -0,032 | 0,032 | -0,189 | -0,197 |
|  | P | 0,472 | 0,121 | **0,015** | 0,340 | 0,866 | 0,867 | 0,317 | 0,297 |
| LV PWd | R | -0,243 | 0,169 | -0,139 | -0,273 | -0,011 | 0,171 | -0,012 | -0,074 |
|  | P | 0,196 | 0,373 | 0,464 | 0,145 | 0,955 | 0,367 | 0,948 | 0,696 |
| IV Ss | R | -0,151 | -0,311 | -0,319 | -0,333 | 0,225 | 0,285 | -0,079 | -0,217 |
|  | P | 0,427 | 0,094 | 0,086 | 0,072 | 0,231 | 0,127 | 0,678 | 0,249 |
| LV IDs | R | -0,220 | -0,005 | -0,410 | -0,182 | -0,064 | -0,094 | -0,002 | -0,223 |
|  | P | 0,242 | 0,979 | **0,025** | 0,336 | 0,735 | 0,622 | 0,993 | 0,236 |
| LV PWs | R | 0,099 | 0,239 | -0,176 | 0,127 | -0,032 | 0,186 | -0,103 | -0,021 |
|  | P | 0,601 | 0,204 | 0,351 | 0,504 | 0,866 | 0,325 | 0,587 | 0,913 |
| LVd mass | R | -0,236 | -0,392 | 0,082 | -0,183 | 0,118 | -0,321 | -0,160 | 0,231 |
|  | P | 0,208 | **0,032** | 0,668 | 0,334 | 0,535 | 0,083 | 0,397 | 0,220 |
| LVs mass | R | 0,215 | -0,492 | -0,094 | -0,128 | 0,290 | 0,005 | -0,013 | -0,004 |
|  | P | 0,253 | **0,006** | 0,621 | 0,501 | 0,121 | 0,979 | 0,945 | 0,981 |

When the systolic and diastolic blood pressure and left ventricular measurements were compared, negative weak correlation was found between systolic blood pressure and LVIDs, while positive moderate correlation was found between systolic blood pressure and LVd mass and LVs mass (Table 5).

**Table 5.** Correlations between blood pressure and left ventricular echocardiography findings.

|  |  | **Sistolic Blood Pressure** | **Diastolic Blood Pressure** |
| --- | --- | --- | --- |
| LVIDd | R | -0,112 | -0,234 |
|  | P | 0,555 | 0,214 |
| LVPWd | R | -0,233 | -0,057 |
|  | P | 0,214 | 0,764 |
| IVSs | R | -0,054 | -0,185 |
|  | P | 0,779 | 0,329 |
| LVIDs | R | -0,373 | -0,057 |
|  | P | **0,042** | 0,765 |
| LVPWs | R | -0,173 | -0,259 |
|  | P | 0,359 | 0,167 |
| LVd mass | R | 0,467 | 0,226 |
|  | P | **0,009** | 0,230 |
| LVs mass | R | 0,543 | 0,222 |
|  | P | **0,002** | 0,239 |
| LVd mass | R | 0,434 | 0,189 |
|  | P | **0,017** | 0,318 |
| LVs mass | R | 0,424 | 0,037 |
|  | P | **0,020** | 0,847 |

When the correlations between inflammation parameters and blood pressure findings were examined, a positive moderate correlation was found between leptin and systolic blood pressure in healthy children, while a negative moderate correlation was found between Pro-BNP and systolic blood pressure and a positive weak correlation was found between leptin and diastolic blood pressure in patient group (Table 6).

**Table 6.** Correlations between blood pressure and inflammation parameters in healthy and obese children.

|  |  | Patient |  | Healty |  |
| --- | --- | --- | --- | --- | --- |
|  |  | Sistolic<br>Blood<br>Pressure | Diastolic<br>Blood<br>Pressure | Sistolic<br>Blood<br>Pressure | Diastolic<br>Blood<br>Pressure |
| CRP | r | 0,057 | 0,275 | 0,098 | -0,011 |
|  | p | 0,763 | 0,141 | 0,607 | 0,955 |
| PRO-BNP | r | -0,566 | -0,114 | -0,108 | 0,066 |
|  | p | **0,001** | 0,547 | 0,571 | 0,728 |
| LEPTİN | r | 0,064 | 0,384 | 0,425 | -0,019 |
|  | p | 0,736 | **0,036** | **0,019** | 0,921 |
| IL-1 | r | -0,161 | -0,301 | -0,272 | -0,011 |
|  | p | 0,395 | 0,106 | 0,147 | 0,955 |
| PENTRAXİN3 | r | **0,037** | 0,054 | 0,187 | 0,136 |
|  | p | 0,846 | 0,777 | 0,323 | 0,475 |

20 of 30 patients evaluated with ABPM. Medical treatment was started for 17 of 20 obese and hypertensive patients in the obesity group, while diet and exercise were recommended to the remaining 3 patients.

Table 7 shows the comparison of the data of patients in the obesity group who were started hypertension treatment and the patients who were not receiving treatment. Leptin, systolic blood pressure, diastolic blood pressure, height, weight and body mass index, IVSd, LVIDd, LVPWd, IVSs, LVIDs, LVPWs and LVs mass values were found to be statistically significantly high in obese patients who were started anti-hypertensive treatment (table 7).

**Table 7.** Comparison of measurements in terms of groups.

|  | GROUP |  |  |  |  |  |  |  |  |  |
| --- | --- | --- | --- | --- | --- | --- | --- | --- | --- | --- |
|  | Obese, treatment started |  |  | Obese, not receiving treatment |  |  | Control |  |  |  |
|  | X | S.S. | Median | X | S.S. | Median | X | S.S. | Median | p |
| Height | 164,00 | 13,78 | 165,00 | 156,85 | 15,02 | 161,00 | 155,64 | 12,76 | 157,50 | 0,041 |
| Weight | 90,66 | 24,26 | 89,00 | 73,87 | 24,50 | 81,00 | 48,47 | 13,04 | 50,50 | 0,000 |
| BMI | 33,02 | 6,44 | 30,86 | 28,92 | 4,79 | 29,02 | 19,62 | 2,85 | 19,56 | 0,000 |
| BMIpercentil | 99,02 | 1,07 | 99,22 | 98,73 | 1,17 | 98,98 | 42,82 | 28,38 | 29,23 | 0,000 |
| SBP | 145,76 | 10,35 | 147,00 | 136,46 | 12,69 | 135,00 | 110,43 | 4,51 | 110,00 | 0,000 |
| DBP | 85,82 | 13,61 | 87,00 | 75,85 | 6,11 | 76,00 | 62,93 | 4,88 | 62,00 | 0,000 |
| CRP | 1,82 | 4,79 | 0,33 | 0,52 | 0,62 | 0,33 | 0,34 | 0,07 | 0,33 | 0,091 |
| PRO-BNP | 33,60 | 21,25 | 24,30 | 72,12 | 81,47 | 41,60 | 47,17 | 32,54 | 38,50 | 0,137 |
| LEPTIN | 15,02 | 12,76 | 7,08 | 13,31 | 8,86 | 14,76 | 2,71 | 3,65 | 1,35 | 0,000 |
| IL-1 | 18018,53 | 74292,30 | 0,00 | 0,00 | 0,00 | 0,00 | 324,90 | 1779,55 | 0,00 | 0,669 |
| PENTRAXIN3 | 75,41 | 84,24 | 49,56 | 60,45 | 21,14 | 51,68 | 66,59 | 57,15 | 51,23 | 0,870 |
| IVSd | 0,60 | 0,13 | 0,56 | 0,52 | 0,12 | 0,54 | 0,46 | 0,12 | 0,45 | 0,006 |
| LVIDd | 2,98 | 0,42 | 2,89 | 2,39 | 0,35 | 2,37 | 2,30 | 0,40 | 2,31 | 0,000 |
| LVPWd | 0,56 | 0,14 | 0,53 | 0,51 | 0,12 | 0,49 | 0,45 | 0,09 | 0,47 | 0,051 |
| IVSs | 0,76 | 0,14 | 0,74 | 0,65 | 0,14 | 0,63 | 0,62 | 0,08 | 0,61 | 0,001 |
| LVIDs | 1,83 | 0,27 | 1,80 | 1,36 | 0,30 | 1,37 | 1,33 | 0,28 | 1,31 | 0,000 |
| LVPWs | 0,92 | 0,25 | 0,85 | 0,76 | 0,16 | 0,77 | 0,74 | 0,14 | 0,73 | 0,009 |
| LVd mass | 93,87 | 38,57 | 80,22 | 80,12 | 16,31 | 78,42 | 77,43 | 23,10 | 75,15 | 0,575 |
| LVs mass | 74,27 | 24,08 | 65,42 | 59,84 | 15,68 | 60,17 | 55,71 | 16,67 | 49,59 | 0,039 |
| LVd mass | 81,60 | 30,80 | 70,16 | 72,26 | 12,92 | 71,31 | 68,58 | 17,74 | 65,92 | 0,482 |
| LVs mass | 62,91 | 19,53 | 56,81 | 61,23 | 13,98 | 57,34 | 50,97 | 13,12 | 48,95 | 0,077 |

## DISCUSSION

Metabolic syndrome (MS) was first described by Reaven in 1988 under the name Syndrome X to show the relationship between insulin resistance and increased risk of lipid disorders, hypertension, type 2 diabetes and atherosclerotic heart diseases in adults. While Metabolic syndrome is known as a health problem mostly seen in adults, it has emerged as an important health problem in children and adolescence in recent years. The increase in the prevalence of obesity in children and adolescents causes an increase in the frequency of Metabolic syndrome. As seen in the pathogenesis of metabolic syndrome in adults, obesity and insulin resistance are mostly held responsible for the pathogenesis in childhood.

Hypertension (HT) and obesity is an important health problem with an increasing prevalance and incidence throughout the world. The number of studies reporting an increase in HT prevalance in childhood in the world are limited. However, regional studies have shown that the prevalance of HT may increase in children over the years. The basis of systemic arterial hypertension, which is an important health problem in adults, starts in childhood and can be detected by blood pressure measurement in childhood (10). Routine blood pressure measurement in hospital referrals in healthy children helps early detection of essential hypertension and early diagnosis of asymptomatic hypertensive patients that develop secondary to other underlying diseases. In studies conducted in different countries, the prevalance of hypertension varies between 1 and 22% (11) (12) (13). The prevalance of hypertension in studies conducted in our country has been reported to be between 3.63 and 17.8% (14).

In their study conducted on 2478 school children between the ages of 12 and 14 in Bursa, Akış et al. found a positive correlation between the prevalance of hypertension and weight gain. In the same study, being overweight was found to be effective in HT prevalance (15). In our study, age, anthropometric measurements, body mass indices and blood pressure measurements were found to be statistically significantly higher in children with obesity.

In their study, Tadahisa N. et al. performed echocardiography on 37 obese patients and compared them in terms of the relationship between left ventricular functions and abdominal fat accumulation. They found that left ventricular end-diastolic volume was significantly high in obese patients when compared with the control group. This study indicated that this increase was directly proportional to body weight. The reason for this has been reported as the increase in fat mass causing hypervolemia and an increase in cardiac output secondary to this. It is thought that the increase in left ventricular diameter increases depending on physiological adaptation (16). In a study conducted on 40 normal individuals and 40 obese patients without secondary damage, Gian F. et al. found that left ventricular end-diastolic volume and left ventricular end-systolic volume was significantly higher in obese patients when compared with healthy children (17).

In the early stage of obesity, left ventricular hypertrophy develops secondary to an increase in blood pressure and left ventricular wall tension due to increased blood volume. Long-term persistence of obesity causes volume and/or pressure load increase, followed by left ventricular dilatation, eccentric hypertrophy, and systolic and diastolic dysfunction. In obese patients, an increase in fatty acid oxidation and myocardial oxygen consumption can be observed due to decreased glucose use secondary to insulin resistance. It is thought that the increase in adipose tissue may affect the conduction system of the heart by causing myocardial atrophy as a result of the pressure on the myocardium, and the lipotoxicity and damage caused by the secreted adipokines and triglyceride increase in the myocytes may cause the deterioration of the systolic function of the heart(18).

In our study, when patient and control group echocardiography findings were compared, intraventricular semptum diastolic diameter (IVSd), left ventricular diastolic diameter (LVIDd), left ventricular free wall diastolic diameter (LVPWd), intraventricular semptum systolic thickness (IVSs), left ventricular systolic diameter (LVIDs) and left ventricular free wall diastolic thickness (LVPWs) were found to be statistically higher in the patient group.

In a study conducted on 49 obese children by Van Putte-Katier et al., left ventricular wall size, especially intraventricular septum diastolic thickness (IVSd), left ventricular diastolic diameter (LVIDs), left ventricular diastolic posterior wall thickness (LPWd) were found to be higher in obese cases when compared with the control group (19). In their study, Erdem A. et al. found that some of the left ventricular measurements of obese children (LVIDs, IVSd and LVdmass) were higher than the control group and especially IVSd was increased in the group which had both HT and insulin resistance (20). In our study, when the correlation between systolic and diastolic blood pressure and left ventricular measurements were examined, we found negative weak correlation between systolic blood pressure and LVIDs (p:0,042 r:-0,373), and positive moderate correlation between systolic blood pressure and LVd mass(p:0,009 r:0,467) and LVs mass (p:0,002 r:0,543); we also found that intraventricular septum diastolic thickness (IVSd), intraventricular septum systolic thickness (IVSs), left ventricular diastolic diameter (LVIDd), left ventricular systolic diameter (LVIDs), left ventricular posterior wall diastolic thickness (LVPWd) and left ventricular posterior wall systolic thickness (LVPWs) were statistically higher in the patient group.

The fact that studies conducted inflammatory cytokines in obesity confirm that obesity is also a chronic inflammatory condition. Insulin resistance and hypertension are a part of obesity-related metabolic syndrome (IRS). Researchers suggest that blood pressure increase is mediated by various bioactive substances secreted from adipose tissue, such as leptin, angiotensinogen, interleukin-6 (IL-6), tumour necrosis factor-α (TNF-α), and adiponectin (21) (22). In a study by Garanty et al. examining serum markers of inflammation and endothelial activation in 50 children with obesity-related hypertension and 143 obese children with normal blood pressure, plasma insulin, glucose, C-reactive protein (CRP), fibrinogen, interleukin-6 (IL-6), interleukin-1β (IL-1β), intercellular adhesion molecule-1 (ICAM-1), vascular cell adhesion molecule-1 (VCAM-1) and lipid levels were evaluated. When children with hypertension were compared with normotensive children, all inflammatory markers and endothelial activation indices were found to be significantly higher. These studies showed that low-grade inflammation and endothelial dysfunction may be closely related to the pathogenesis of obesity-associated hypertension at early age (23). When the correlations between inflammation parameters and blood pressure findings were examined in our study, positive and moderate (p:0,019 r:0,425) correlation was found between leptin and systolic blood pressure in healthy children, while negative moderate (p:0,001 r:-0,566) correlation between Pro-BNP and systolic blood pressure and positive weak correlation between leptin and diastolic blood pressure (p:0,036 r:0,384) were found in obese and hypertensive children.

In Our study, statisticaly significant correlation was found between leptin and echocardiography results of the patient and control group. Negative moderate correlation was found between left ventricular diastolic mass (**LVd mass)** (p:0,032 r:-0,392) and left ventricular systolic mass (**LVs mass**) (p:0,006 r:-0,492) and Pro-BNP in the patient group. Negative weak correlation was found between left ventricular diastolic diameter (**LVIDd)** (p:0,015 r:-0,441), left ventricular systolic diameter (**LVIDs)** (p:0,025 r:-0,410) and Pro-BNP in the patient group. When the correlation between Pentraxin-3 and echocardiographic measurements was evaluated, no statistically significant correlation was found.

In previously studies and also in our study, inflammatory markers were found to be higher in obese individuals with hypertension. It can be seen that inflammation caused by obesity is an important determining factor for hypertension. In our study, a moderate positive correlation was found between leptin and pent-raxin-3 in obese children (p:0.023, r:0,413). In terms of the correlation between inflammation parameters and blood pressure findings, moderate positive correlation was found between leptin and systolic blood pressure in healthy children, while moderate negative correlation (p:0.001, r:-0.566) between Pro-BNP and systolic blood pressure and weak positive correlation (p:0.036, r:0.384) between leptin and diastolic blood pressure were found in patient group.

In the study by Duprez et al., it was shown that there is a significant relationship between CRP and early atherosclerosis. Hommels et al. showed a moderate relationship between CRP and abdominal aortic and renal artery atherosclerosis. Taken together, Duprez et al. and Hommels et al.’s studies support the hypothesis that the vascular effects of inflammation in the walls of the great arteries (manifested by increased hsCRP levels) are due to decreased elasticity and increased stiffness of the large arteries, which is one of the markers of development of atherosclerosis. The proinflammatory process, which starts with mediators such as adhesion molecules, chemokines, growth factors, heat shock proteins, endothelin-1 and angiotensin, shows that inflammation may play a role in the initiation and progression of hypertension. Increased cytokines may impair the capacity to produce endothelium-derived vasodilator factors, especially nitric oxide (NO), resulting in endothelial dysfunction, chronic impaired vasodilation, and hypertension.(24,25)

Inflammation has an important role in the occurrence and prognosis of cardiovascular diseases. Hypertension is one of the most frequent systemic diseases. Inflammation has an important role in the pathogenesis of hypertension and contributes to the formation of target organ damage due to hypertension (26).

In a study conducted by Wang et al., it was found that losing weight with a four-week long healthy diet and exercise caused a significant decrease in inflammation in overweight adolescents (27). Patients with high inflammation parameters may need more medical treatment in addition to lifestyle changes. In our study, leptin, systolic blood pressure, diastolic blood pressure, height, weight and body mass index, IVSd, LVIDd, LVPWd, IVSs, LVIDs, LVPWs and LVs mass values were found to be statistically significantly high in obese patients who were started anti-hypertensive treatment.

The aim of our study is evaluate levels of CRP, IL-1, proBNP, leptin and PTX-3 to evaluate inflammation in children with obesity-related hypertension. When measurements were compared with patient and control group, CRP and leptin values were found to be statistically significantly high in patient group. No significant difference was found between groups in Pentraxin-3, IL-1 and Pro-BNP levels. Early diagnosis and treatment are important to prevent hypertension complications. In our study, we found that CRP and leptin values, which are the markers we examined for the evaluation of inflammation, were high in obese and hypertensive patients. We think that the high levels of inlammation markers in obese patients may be a guide in the severity of hypertension, in the prevention of organ damage secondary to hypertension, in reversing inflammation and hypertension with more stringent lifestyle changes, diet and exercise.

In conclusion, when the parameters in the patient and control groups were compared, it was found that CRP and leptin values were statistically significantly higher in the patient group, and there was no significant difference between the groups in Pentraxin-3, IL-1 and Pro-BNP values. We think that CRP and leptin play a role in the pathogenesis of obesity-associated early hypertension and that these parameters may be a guide in reversing inflammation and hypertension with more strict lifestyle changes, diet and exercise.

We think that the high levels of inlammation markers in obese patients may be a guide in the severity of hypertension, in the prevention of organ damage secondary to hypertension, in reversing inflammation and hypertension with more stringent lifestyle changes, diet and exercise.

The severity of hypertension being an inflammatory disease may guide the development of new therapeutic strategies to reduce the morbidity and mortality of hypertension and to treat hypertension-related target organ damage. However, prospective studies are needed to evaluate the effects and prognosis of these interventions.

## Data Availability

All relevant data are within the manuscript and its Supporting Information files.

